# Advancing in Schaaf-Yang syndrome pathophysiology: from bedside to subcellular analyses of truncated MAGEL2

**DOI:** 10.1101/2022.05.04.22274475

**Authors:** Laura Castilla-Vallmanya, Mónica Centeno-Pla, Mercedes Serrano, Héctor Franco-Valls, Raúl Martínez-Cabrera, Aina Prat-Planas, Elena Rojano, Juan A. G. Ranea, Pedro Seoane, Clara Oliva, Abraham J. Paredes-Fuentes, Rafael Artuch, Daniel Grinberg, Raquel Rabionet, Susanna Balcells, Roser Urreizti

**Author notes:** These authors contribute equally (Laura Castilla-Vallmanya, Mónica Centeno-Pla, Mercedes Serrano, Raquel Rabionet, Susanna Balcells, Roser Urreizti). **Corresponding author:** Dr. Roser Urreizti, Passeig de Sant Joan de Déu, 2, 08950 Esplugues de Llobregat, Barcelona.

## Abstract

**Background:** Schaaf-Yang syndrome (SYS) is caused by truncating mutations in *MAGEL2*, mapping to the Prader-Willi region (15q11-q13), with an observed phenotype partially overlapping that of Prader-Willi syndrome. MAGEL2 plays a role in retrograde transport and protein recycling regulation. Our aim is to contribute to the characterization of SYS pathophysiology at clinical, genetic and molecular levels.

**Methods:** We performed an extensive phenotypic and mutational revision of previously reported SYS patients. We analysed the secretion levels of amyloid-β 1-40 peptide (Aβ_1-40_), and performed targeted metabolomic and transcriptomic profiles in SYS patients’ fibroblasts (n=7) compared to controls (n=11). We also transfected cell lines with vectors encoding wild-type (WT) or truncated MAGEL2 to assess stability and subcellular localization of the truncated protein.

**Results:** Functional studies show significantly decreased levels of secreted Aβ_1-40_ and intracellular glutamine in SYS fibroblasts compared to wild-type. We also identified 132 differentially expressed genes, including ncRNAs such as HOTAIR, many of them related to developmental processes and mitotic mechanisms. The truncated form of MAGEL2 displayed a stability similar to the wild-type but it was significantly switched to the nucleus, compared to a mainly cytoplasmic distribution of the wild-type MAGEL2. Based on updated knowledge we offer guidelines for clinical management of SYS patients.

**Conclusion:** A truncated MAGEL2 protein is stable and localises mainly in the nucleus, where it might exert a pathogenic gain of function effect. Aβ_1-40_ secretion levels and HOTAIR mRNA levels might be promising biomarkers for SYS. Our findings may improve SYS understanding and clinical management.

**Key Messages:** *MAGEL2* truncating mutations cause Schaaf-Yang syndrome (SYS) but the functional effects of the truncated MAGEL2 protein have been poorly defined. By expressing truncated MAGEL2 in cell lines, we now know that a truncated version of the protein is retained in the nucleus, thus exerting a gain-of-function behaviour in addition to the loss of some of its main functions. Patients’ fibroblasts show reduced levels of excreted amyloid beta 1-40 and intracellular glutamine as well as an altered transcriptomic profile, including overexpression of the major regulator HOTAIR. Based on a comprehensive review of genetic and clinical aspects of all reported cases, families and physicians will benefit from the Clinical Management Recommendations that we provide here.

## INTRODUCTION

In 2013, truncating mutations in *MAGEL2* (OMIM* 605283) were associated with a new clinical entity,**[1]** first described as a Prader-Willi-like syndrome and currently named Schaaf-Yang syndrome (SYS; OMIM# 615547). *MAGEL2* is one of the five maternally imprinted protein-coding genes contained in the Prader-Willi region (15q11-q13). Lack of expression of the paternal alleles in this region causes Prader-Willi syndrome (PWS; OMIM #176270). In contrast, nonsense or frameshift mutations in the paternal allele of *MAGEL2* are predicted to encode a truncated protein lacking the MAGE Homology Domain (MHD) domain and have been associated with SYS.**[1]** Since then, over a hundred SYS patients have been reported and phenotypically described.**[1–28]** SYS patients and PWS patients show overlapping clinical phenotypes, including neonatal hypotonia, intellectual disability (ID), developmental delay (DD), early feeding difficulties, endocrinologic disturbances (hypogonadism and other hormonal dysbalances) and sleep disorders. However, some of the clinical criteria for PWS diagnosis, such as hypopigmentation, characteristic facial dysmorphisms, small hand and feet, hyperphagia, obesity and obsessive-compulsive behaviours, are frequently absent in SYS patients, who, on the other hand, present more frequently with severe ID, autism spectrum disorder (ASD) behaviours and joint contractures.**[29–31]** Some truncating variants in *MAGEL2* have also been associated with Chitayat-Hall syndrome (CHS).**[32]** However, a systematic review of all SYS and CHS patients showed that there is no discernible genetic or clinical difference between both syndromes, and the latter has been renamed as SYS in OMIM.**[17]** In contrast, two particular *MAGEL2* truncating mutations have been recurrently identified in patients affected by lethal arthrogryposis multiplex congenita (AMC), a much more severe phenotype, distinct from SYS.**[4, 10, 27, 33, 34]** All in all, there is no specific constellation of symptoms pathognomonic or specific for any of these clinical syndromes; furthermore, they probably conform to a clinical continuum, therefore denoting the need to address clinical denomination according to molecular findings.

*MAGEL2* shows a wider expression in human foetal tissues than in adult tissues, where it is predominantly present in brain (according to GTEx **[35]**). In adult mice, it also becomes mostly restricted to the central nervous system, specifically to the amygdala and the hypothalamus, and predominantly in the suprachiasmatic, the paraventricular and the supraoptic nuclei.**[36–38]** It is a single-exon gene that encodes one of the largest proteins of the Type II MAGE protein family, of 1249 amino acids. At a structural level, the N-terminal region of MAGEL2 contains a proline-rich domain, whose function remains unclear.**[39]** At the C-terminus, from amino acid 1027 to 1195, there is the MHD, a highly conserved 170-amino acid sequence present in both type I and type II MAGEs, crucial for protein-protein interaction.**[40]** Through it, MAGEL2 recognizes and binds the coiled-coil domain of the E3 ubiquitin ligase TRIM27. The MHD is also crucial for binding VPS35, a subunit of the retromer cargo-selective complex. MAGEL2, TRIM27 and USP7 form the MUST complex, which is recruited to endosomes through direct binding of MAGEL2 to VPS35 and plays a role in retrograde endosomal transport.**[41, 42]** These specialised endosomes participate in endosomal export pathways that deliver membrane protein cargoes either to the trans-Golgi network through retrograde pathways or to the plasma membrane through recycling pathways.**[43]**

A dysfunction of the retrograde transport could be disturbing for many cellular processes. Loss of *MAGEL2* expression causes a reduction in secretory granules protein levels due to impaired endosomal protein trafficking and subsequent lysosomal degradation, resulting in a reduction of circulating bioactive hypothalamic hormones.**[36]** A well-coordinated trafficking network is also key for the correct regulation of amyloid precursor protein (APP) cleavage.**[44]** APP family members are relevant for neuronal differentiation and migration during cortical development **[45, 46]** and proper neuromuscular junction formation and neurotransmission.**[47]** Many studies support a model where retromer deficiency leads to increased APP cleavage, Aβ peptide production and exocytosis.**[48–50]** In addition, protein levels of the glucose transporter GLUT1 in the cell membrane are reduced after VPS35 and SNX27 inhibition, showing that they are also tightly regulated by the retromer.**[51]**

Here, we have performed an extensive literature revision and based on it, we have expanded the clinical and genetic delineation of SYS and developed a standardised set of guidelines for its clinical management. We also contribute to the knowledge of the cellular phenotype by assessing the effect of a recurrent truncating variant on MAGEL2 protein stability and subcellular localization using heterologous expression vectors. Finally, we have performed a transcriptomic and metabolomic characterization of fibroblasts derived from SYS patients, and interpreted the results in the context of molecular and clinical findings.

## RESULTS

### Clinical management of SYS patients requires a coordinated multidisciplinary approach

We performed a systematic revision of all the published patients with SYS to date, who were all carriers of MAGEL2 mutations (**Table 1** and **Supplementary Table 1**), carefully inspecting both the molecular and phenotypic data, with the aim to propose a set of guidelines for SYS clinical management.

**Table 1.**
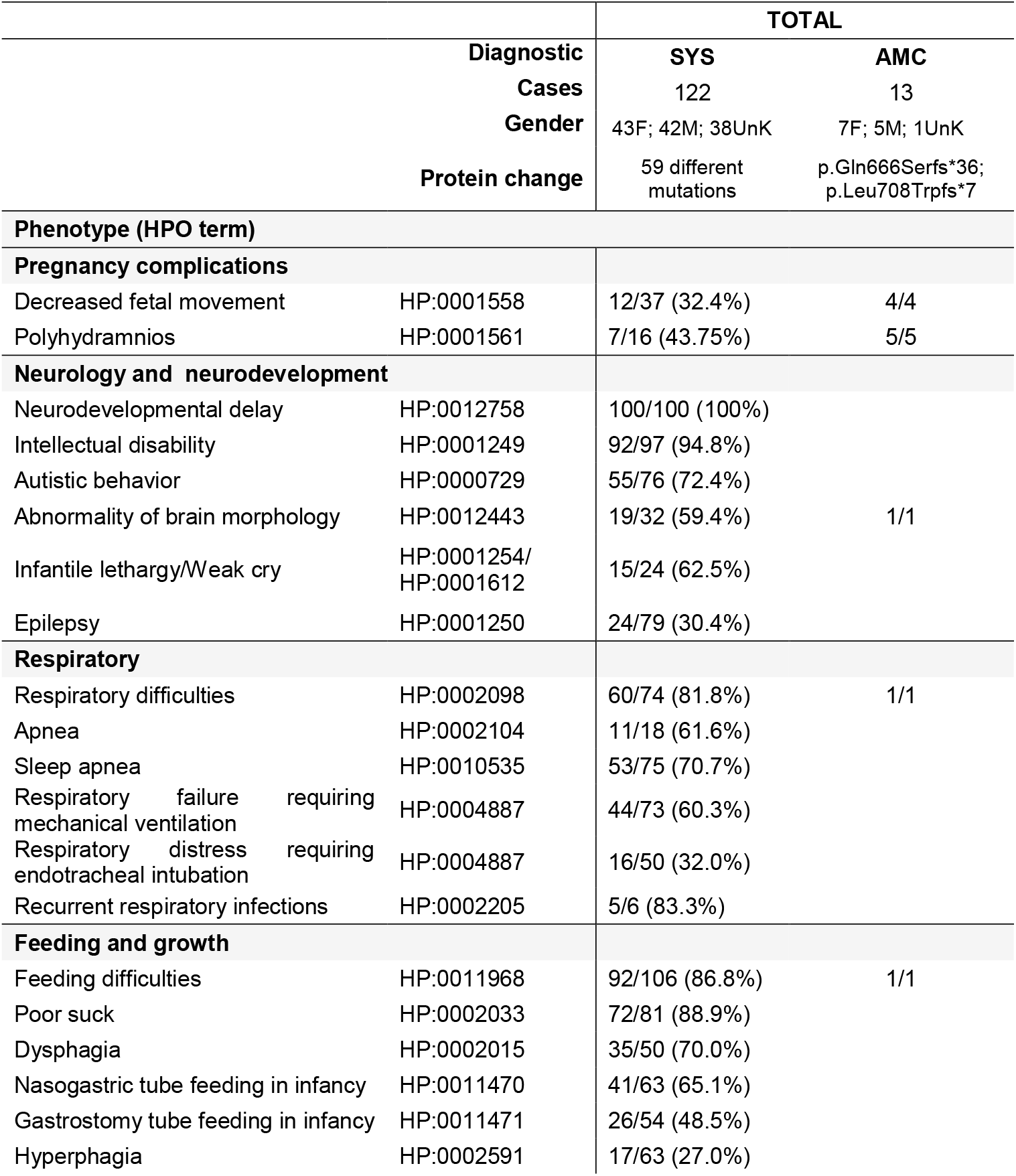
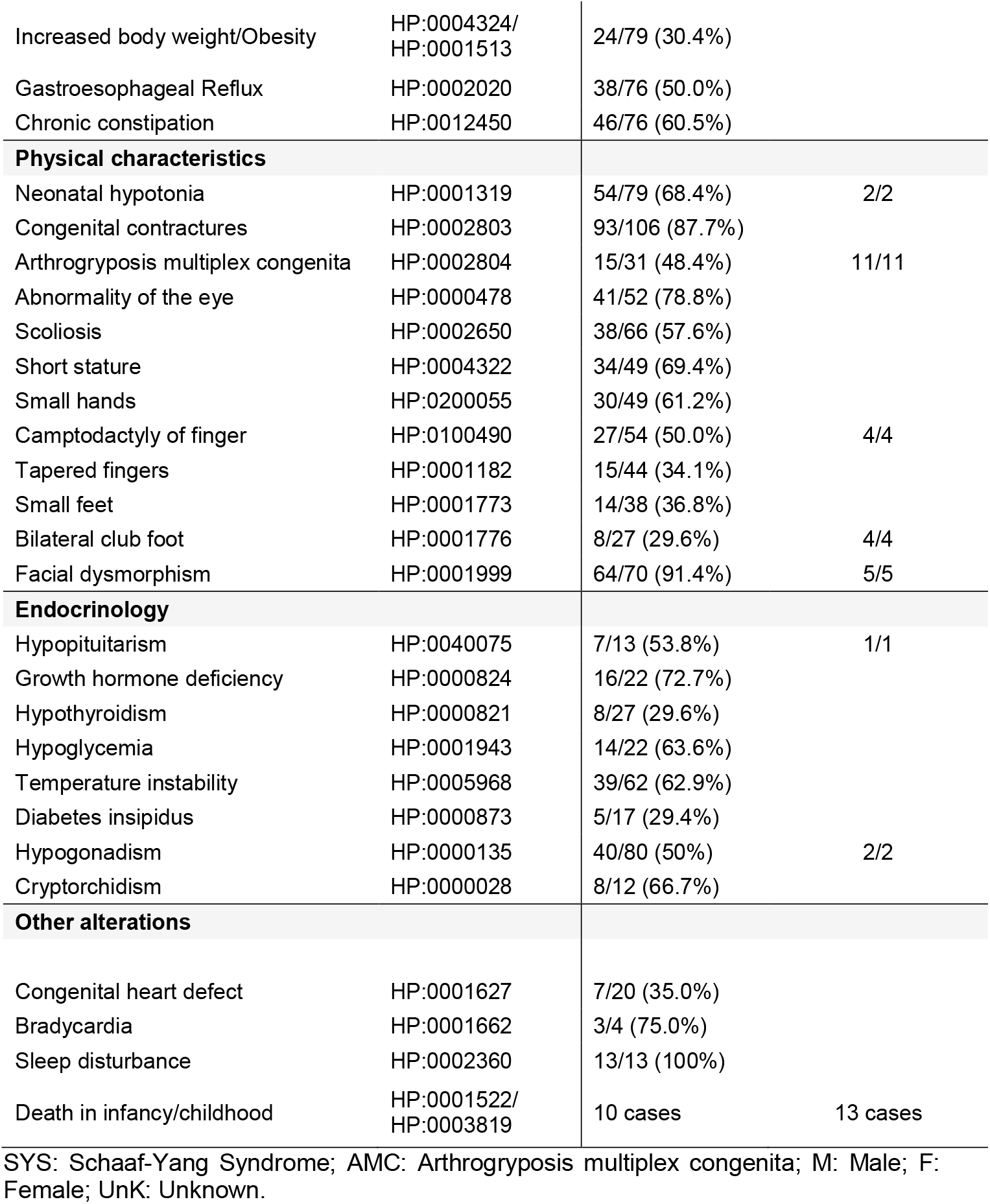
Clinical overview of patients with MAGEL2-related disorders

The literature-based recommendations have been divided into two life periods: the perinatal period (first 28 days of life) and infancy/adolescence. They include the most relevant medical problems associated with each period and the specific concerns or interventions advisable for each area. A schematic version of the different clinical areas and tests included in the guidelines are represented in **Figure 1** and a detailed, printable version is supplied as **Supplementary Table 2** (available In Spanish in **Supplementary Table 3**).

**Figure 1.**
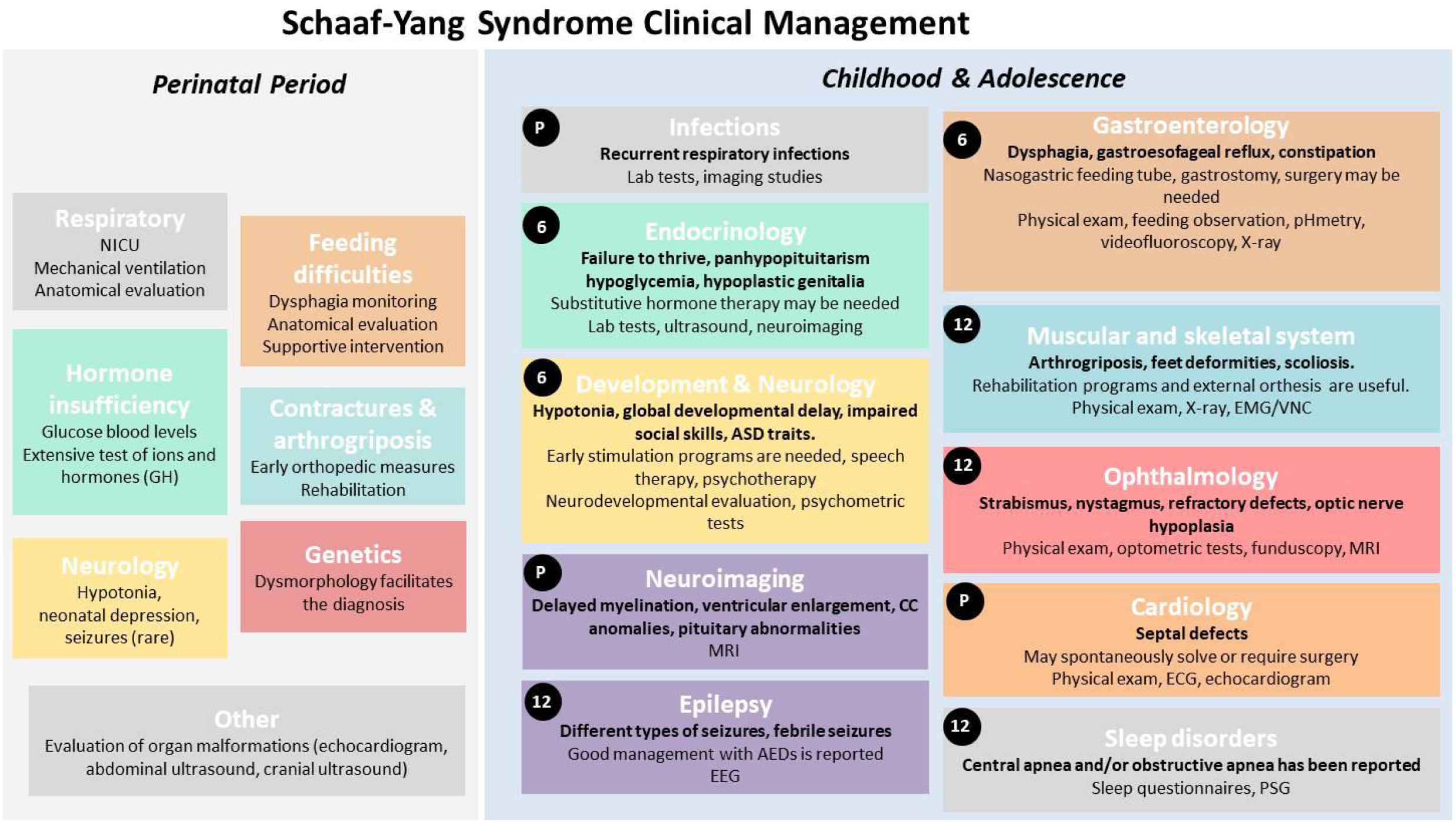
Schematic guidelines for Schaaf-Yang syndrome clinical management. NICU: neonatal intensive care unit; ASD: Autism spectrum disorder; ECG: electrocardiogram; EEG: Electroencephalogram; EMG/NCV: electromyogram/nerve conduction velocity; MRI: Magnetic resonance imaging; PSG: Polysomnography. Boxes indicate the medical area of disease. Medical problems during childhood and adolescence are highlighted in bold. Numbers in black dots state the recommended periodicity for clinical evaluation in months for every specialty. P means personalised, based on comorbidities. Periodicity of Cardiology and Neuroimaging evaluations should be patient-centred. See Supplementary Table 2 for details. A Spanish version of this figure is available on demand.

In most instances, pregnancy was uneventful (only polyhydramnios has been reported in some cases), although there is a high rate of caesarean sections (over 50% of the patients where delivery information is available). Clinical symptoms appear early in life showing a complex phenotype that includes neuromuscular symptoms, respiratory and endocrinological problems, feeding difficulties, and dysmorphic traits. Other clinical issues can appear later and may affect almost any organ or system, requiring a coordinated multidisciplinary approach.

### SYS variants are mostly truncating and located in the C-terminal domain

To date, 61 different variants have been associated with SYS or AMC (**Figure 2**). Mutation p.(Gln666Profs*47), present in 80 individuals, is considered a recurrent mutational hotspot. These variants are mostly truncating and predominantly located in the C-terminal region of the protein, leading to a partial or a total lack of the MHD domain, and compromising the functions that MAGEL2 carries out through this domain. Three different missense variants and a small in-frame duplication have also been associated with SYS phenotypes (in grey in **Figure 2**). These atypical variants are not predicted to encode a truncated form of the protein, and there are no functional studies to support their pathogenicity. Thus, their clinical implication remains to be proven.

**Figure 2.**
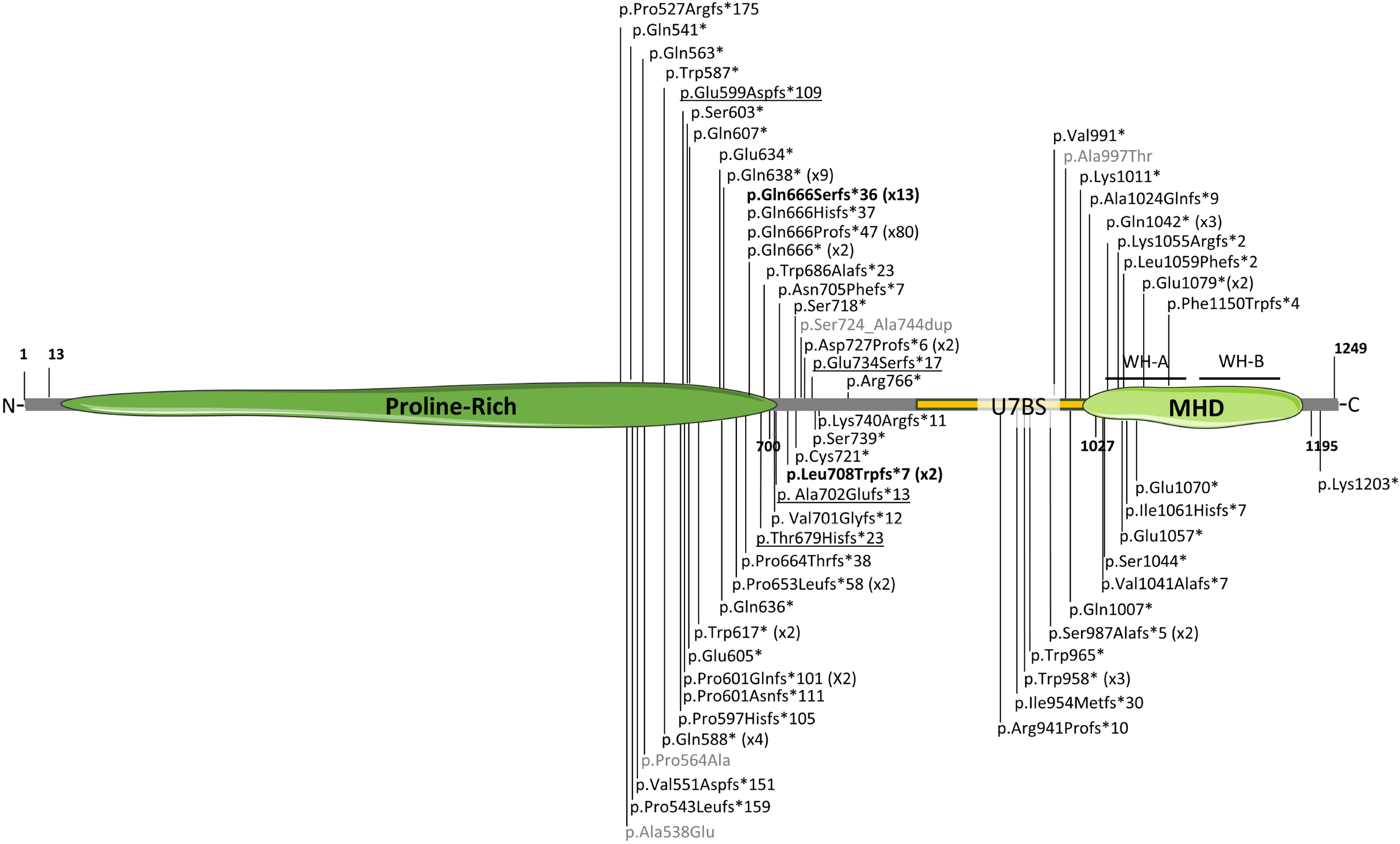
Schematic representation of MAGEL2 disease associated variants. The number in brackets indicates the number of individuals carrying recurrent mutations. In bold, mutations p.Gln666Serfs*36 (x13) and p.Leu708Trpfs*7 (x2) associated with AMC. In grey, atypical, non-truncating variants. Previously misreported variants c.1797_1820del (p.Glu599Aspfs*109 (c.1797_1810del)), c.224delC (p.Thyr679Hisfs*23 (c.2034delC)), c.2015delC (p.Ala702Glufs*13 (c.2105delC)) and c.390delA (p.Glu734Serfs*17 (c.2199delA))**[10]** are underlined. All variants are referenced on transcript NM_019066.5. The complete list of variants, their cDNA annotation and reference are collected in **Supplementary Table 3**. Image created with Servier Medical Art (smart.servier.com).

### Fibroblasts from SYS patients show altered gene expression patterns

To better understand the effect of *MAGEL2* truncating mutations on gene expression patterns, we performed an mRNA whole transcriptome analysis (mRNASeq) on fibroblasts from six healthy donors and three SYS subjects: a girl carrying p.(Gln638*) and a boy and an unrelated girl, both carrying p.(Gln666Profs*47). Using the ExpHunter Suite, we identified 132 differentially expressed genes (DEGs), 76 up-regulated and 56 down-regulated (**Supplementary Table 3**). The top ten up- and down-regulated genes, showing the most significant changes in the expression fold, are listed in **Table 2**. Four genes were tested by qPCR in fibroblasts from 5 SYS patients and 4 control individuals, which confirmed significant upregulation of *HOTAIR* and *PITX1* and downregulation of *TBX5* (**Supplementary Figure 1**).

**Table 2.**
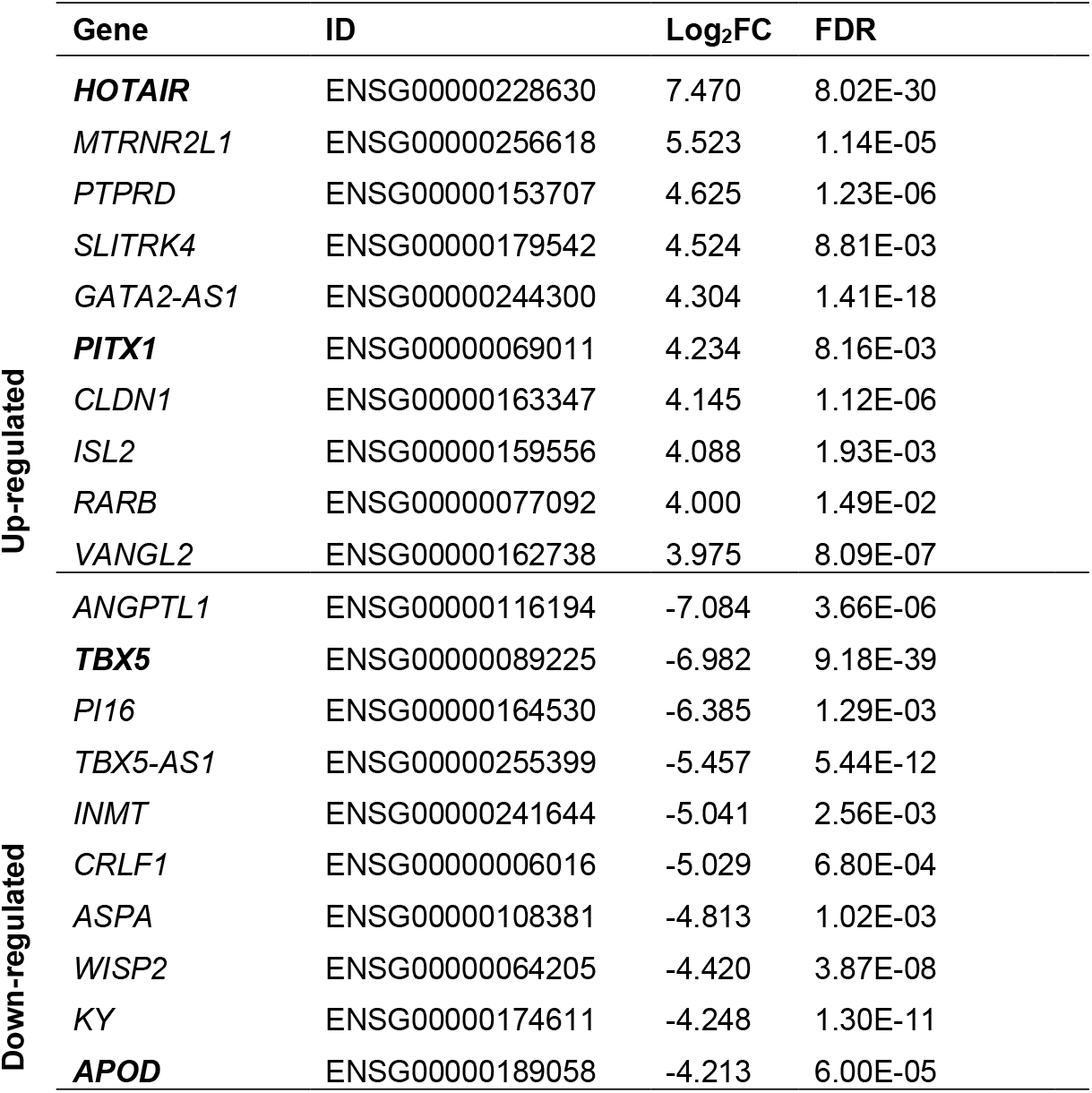
Top ten up- and down-regulated DEGs identified after mRNASeq analysis of skin fibroblasts in SYS patients and controls. In bold, genes analysed by qPCR.

|  | Gene | ID | Log<sub>2</sub>FC | FDR |
| --- | --- | --- | --- | --- |
| Up-regulated | **HOTAIR** | ENSG00000228630 | 7.470 | 8.02E-30 |
|  | *MTRNR2L1* | ENSG00000256618 | 5.523 | 1.14E-05 |
|  | *PTPRD* | ENSG00000153707 | 4.625 | 1.23E-06 |
|  | *SLITRK4* | ENSG00000179542 | 4.524 | 8.81E-03 |
|  | *GATA2-AS1* | ENSG00000244300 | 4.304 | 1.41E-18 |
|  | **PITX1** | ENSG00000069011 | 4.234 | 8.16E-03 |
|  | *CLDN1* | ENSG00000163347 | 4.145 | 1.12E-06 |
|  | *ISL2* | ENSG00000159556 | 4.088 | 1.93E-03 |
|  | *RARB* | ENSG00000077092 | 4.000 | 1.49E-02 |
|  | *VANGL2* | ENSG00000162738 | 3.975 | 8.09E-07 |
| Down-regulated | *ANGPTL1* | ENSG00000116194 | -7.084 | 3.66E-06 |
|  | **TBX5** | ENSG00000089225 | -6.982 | 9.18E-39 |
|  | *PI16* | ENSG00000164530 | -6.385 | 1.29E-03 |
|  | *TBX5-AS1* | ENSG00000255399 | -5.457 | 5.44E-12 |
|  | *INMT* | ENSG00000241644 | -5.041 | 2.56E-03 |
|  | *CRLF1* | ENSG00000006016 | -5.029 | 6.80E-04 |
|  | *ASPA* | ENSG00000108381 | -4.813 | 1.02E-03 |
|  | *WISP2* | ENSG00000064205 | -4.420 | 3.87E-08 |
|  | *KY* | ENSG00000174611 | -4.248 | 1.30E-11 |
|  | **APOD** | ENSG00000189058 | -4.213 | 6.00E-05 |

Enrichment analysis on the 132 identified DEGs highlighted a group of 5 genes related to “collagen formation” and various mitosis related REACTOME categories, such as “Resolution of Sister Chromatid Cohesion” and “Mitotic Spindle Checkpoint”. Consistently, most of those genes were also present in the “Kinetochore” and “Chromosome, centromeric region” categories according to GO cellular components enrichment (**Supplementary Table 4**).

### SYS fibroblasts show decreased Aβ_1-40_ peptide secretion levels

As alterations in *MAGEL2* could be affecting APP cleavage and Aβ_1-40_ peptide production rates, levels of Aβ_1-40_ and Aβ_1-42_ peptides were measured by ELISA in extracellular medium from SYS, PWS and control fibroblasts. SYS fibroblasts showed significantly decreased extracellular levels of the Aβ_1-40_ processed peptide, both compared to PWS and control fibroblasts. No differences were observed in the PWS group compared to the control (**Figure 3A**). Levels of Aβ_1-42_ peptide were extremely low and no differences were observed between conditions (data not shown).

**Figure 3.**
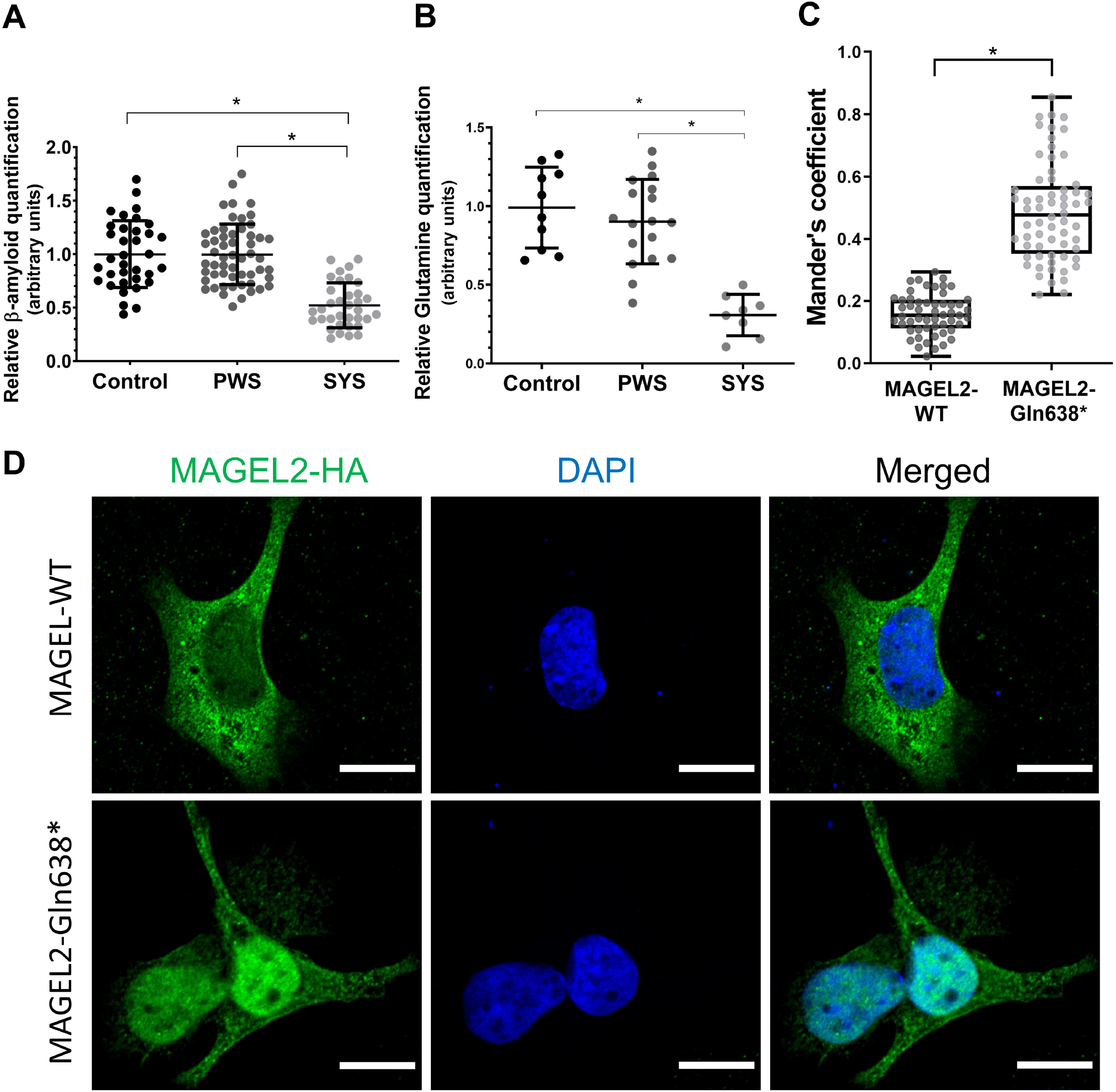
Molecular and cellular biomarkers for SYS. **A)** Aβ_1-40_ peptide levels in control, PWS and SYS fibroblasts extracellular medium. Data obtained from at least 3 independent experiments (SYS: n=5; PWS: n=9; control: n=5). **B)** Glutamine levels in control, PWS and SYS fibroblasts (SYS: n=6; PWS: n=9; control: n=6). Values from 2 independent experiments have been normalised to the mean of the control group. Horizontal lines represent mean values and error bars represent the standard deviation. Statistical analyses were performed using One-Way ANOVA and Tukey’s multiple comparisons test in GraphPad Prism. \**p*< 0.001. **C)** Mander’s coefficient between the HA signal and DAPI in MAGEL2-WT and MAGEL2-Gln638* transfected cells. *n*= 118 from 6 independent experiments. \**p*< 0.001. **D)** Representative immunofluorescence images of SAOS-2 cells transfected with MAGEL2-WT and MAGEL2-Gln638* plasmids, stained with anti-hemagglutinin (HA; green, tagging MAGEL2) or DAPI (blue, cell nuclei). Scale bar represents 15 μm.

### SYS fibroblasts show altered levels of organic acids and amino acids (metabolic profiling in fibroblasts)

Mass spectrometry analysis of intracellular metabolites from extracts of SYS (n=4), PWS (n=9) and control (n=5) fibroblasts showed a robust and significant decrease in glutamine levels for SYS fibroblasts (**Figure 3B**), as well as a significant increase of suberic, sebacic, adipic and malic organic acids levels (**Supplementary Figure 2**).

We investigated if any of the genes involved in these metabolites’ pathways were differentially expressed in the transcriptomic analysis (**Supplementary Table 3**). The only differentially expressed gene involved in glutamine (GO:0006541) or organic acid (GO:0006082) metabolic processes was *ME1* (ENSG00000065833), encoding the NADP-dependent malic enzyme protein, which was upregulated in SYS fibroblasts.

### A truncated form of MAGEL2 remains stable in the cell

To evaluate the stability and recycling of a truncated form of MAGEL2, HEK293T cells were transfected with hemagglutinin (HA)-tagged expression vectors containing either the wild-type (WT) cDNA sequence of *MAGEL2* (MAGEL2-WT) or the c.1912C>T; p.Gln638* (MAGEL2-Gln638*) mutation, which encodes a protein 611 amino acids shorter than the WT form and has been reported in nine patients (Supplementary Table 1). Blocking of the proteasomal degradation pathway with MG132 (**Supplementary Figure 3, A**) or the lysosomal pathway with bafilomycin (**Supplementary Figure 3, B**) showed that MAGEL2 is mostly degraded via proteasome with no differences between WT and truncated MAGEL2 (**Supplementary Figure 3, A and B**). A time-course of cycloheximide treatment (2, 4, 6, 12h) showed also a similar stability and half-life for both MAGEL2 forms (**Supplementary Figure 3, C**).

### A truncated form of MAGEL2 shifts to a nuclear localization

To determine the subcellular localisation of the p.Gln638* truncated form of MAGEL2, HEK293T, SAOS-2 and HeLa cells were transfected with the expression vectors MAGEL2-WT or MAGEL2-Gln638*. Immunocytochemistry assays detecting HA showed that in SAOS-2 cells transfected with the MAGEL2-Gln638* construct, there was a shift of the protein towards the nucleus, while the MAGEL2-WT form was mainly located in the cytoplasm (**Figure 3 C and D**). A similar localisation pattern was observed in HEK293T and HeLa cells (data not shown), supporting the idea that presence of variant p.Gln638*, which leads to the lack of part of the C-terminal sequence, affects protein subcellular localisation independently of the cell type. This result is consistent with predictions obtained following the Scandinavian Protocol, which combines different online tools to predict protein subcellular localisation.**[52, 53]**

## DISCUSSION

Since its first description in 2013,**[1]** more than 150 SYS individuals carrying MAGEL2 truncating mutations have been published. Most publications include one or a reduced group of patients, often with scarce clinical descriptions, hampering the definition of the syndrome’s characteristics. Families, already burdened by the solitude of having an ultra-rare condition, lack clear and established clinical guidelines for their treatment and follow-up. Once available, such evidence-based recommendations will help reduce inequity in health care and empower both families and clinicians facing such a rare disease. To develop the recommendations, we performed an extensive revision of all the SYS subjects published so far at both the phenotypic and genetic level and elaborated a comprehensive and detailed follow-up programme. Despite the comprehensive revision, to date, no clear underlying phenotype-genotype correlation was observed, with the exception of two particular variants [p.(Gln666Serfs*36) and p.(Leu708Trpfs*7)] which are associated with the much more severe phenotype of AMC leading to perinatal death.

Genetically, SYS is caused by nonsense or frameshift mutations. Only one *MAGEL2* missense variant (c.1613C>A; p.Ala538Glu) has been reliably described as potentially disease-associated.**[17]** While at the moment of its publication this change was classified as “disease causing” based on available *in silico* and frequency data, currently, updated information, including a gnomAD V3.1 Amish MAF of 0.03, supports its re-classification as “likely benign” according to ACMG guidelines.**[54]** Clinically, the patient carrying this variant presented with DD, ASD, and dysmorphic traits. While her presentation shares some traits with SYS, the high frequency of this variant in the Amish population, together with the nonspecific character of these traits, would suggest that it is not the main cause of the disease. Two other missense mutations have been identified in two different patients: p.Pro564Ala and p.Ala997Thr (with two and four carriers in gnomAD v3.1.1, respectively). However, their potential causality was not further discussed and there are no clinical descriptions of the patients.**[10]**. An in-frame duplication in the paternal chromosome (p.Ser724_Ala744dup, two carriers in gnomAD v.3.1) has also been identified in a patient with a clinical presentation sharing some resemblances with SYS, but the scarcity of available information makes it difficult to fully understand the clinical role of this variant.**[21]** We have also noticed some missannotations, probably due to the fact that, originally, MAGEL2 was predicted to encode a 529 amino-acid protein (instead of the current 1249 amino acids) lacking the N-terminal domain (hg38). This is the case for reported variants p.(Thr76Hisfs*23) and p.(Glu131Serfs*17),**[10]** which we have relabelled as p.(Thr679Hisfs*23) and p.(Glu734Serfs*17). To sum up, while the pathogenicity of MAGEL2-truncating mutations has been clearly established and documented, it remains to be clarified whether any MAGEL2 missense or in-frame mutation is really a causal variant for SYS.

The phenotypic overlap between SYS and PWS suggests that the alteration of MAGEL2 may contribute to some aspects of the PWS phenotype, but the extent of this contribution is still an open question. Two patients carrying atypical deletions involving only *MAGEL2, NDN* and *MKRN3*, did not show a full PWS phenotype: one patient showed only mild delayed motor skills and the other displayed obesity, DD and high pain threshold.**[55, 56]** At the same time, neither does the deletion lead to SYS, further supporting that it is the presence of the truncated form of the MAGEL2 protein that leads to some of the particular aspects of SYS. As our results show, the truncated protein is synthesised, it is stable and it is not degraded any faster than the WT form. We propose that this truncated form could be exerting toxic effects in addition to the negative effects caused by the lack of WT MAGEL2.

The role of MAGEL2 in the regulation of endosomal protein trafficking and recycling has been widely studied.**[41, 42, 57]** Consistent with this, loss of paternal MAGEL2 expression in Magel2^pΔ/m+^ mice and PWS patient–derived neuronal cell models, leads to decreased levels of secretory granule proteins, which lead to reduced levels of circulating bioactive hormones and of mature secretory granules in neurons.**[36]** Also, dental pulp stem cell derived neurons from several PWS patients and one SYS patient showed impaired trafficking of M6PR (cation-dependent mannose-6-phosphate receptor), indicating impaired endosome-mediated retrograde transport. We hypothesised that this impaired trafficking could also be reflected in an aberrant Aβ peptide production and exocytosis.**[48–50]** Indeed, Aβ_1-40_ peptide extracellular levels of SYS fibroblasts were significantly decreased compared to those of PWS and control groups, which showed no difference between them. This result supports the idea that the truncated form of the protein may have a different effect on protein trafficking than the complete loss of the protein itself and establishes Aβ_1-40_ levels as a potential biomarker for the disease in patient-derived cells.

Despite being its most deeply studied function, dysregulation of protein trafficking and recycling is not the only consequence of MAGEL2 malfunction. The subcellular localisation of the full MAGEL2 protein **[41, 42]** and its C-terminal part alone,**[39]** have been previously studied in heterologous systems, both presenting a mainly cytoplasmic localisation. Our immunocytochemistry assays in different transfected cell lines showed that, in contrast, the heterologously expressed MAGEL2-Gln638* (N-terminal part of the protein) was predominantly located inside the nucleus. The new localisation of the protein suggests that, in addition to the loss of the MAGEL2 normal functions, including the translocation of YTHDF2 to the nucleus,**[39]** SYS-associated mutations could involve new functions of unknown consequences.

To explore if the truncated form of MAGEL2 could be affecting the expression of other genes, we applied an unbiased transcriptomic approach in fibroblasts. We found an enrichment in genes related to mitosis, nuclear function and localisation, as well as in developmental and neurologic processes.

Previous studies on the full length and C-terminal parts of the MAGEL2 protein have shown that this protein is involved in RNA metabolic processes.**[39]** In our transcriptomic analysis, *HOTAIR* expression was clearly increased in SYS fibroblasts. *HOTAIR* is a well-known long non-coding RNA that mediates transcriptional silencing in *trans*. This gene’s promoter contains binding sites for numerous transcription factors, including AP1, Sp1 and NF-κβ.**[58]** It is able to exert epigenetic functions through H3K27 trimethylation and H3K4 demethylation, among other regulatory functions, and has a widely studied role in cancer (reviewed in **[59]**). HOTAIR promotes transcriptional silencing of the *HOXD* locus, which plays an essential role in determining body axes and orchestrating organ formation during vertebrate development.**[60]** HOTAIR is also a regulator of mTOR, increasing its phosphorylation and mTOR mediated exosome release,**[61]** and its regulation of GLUT1 levels (**Figure 4**).**[62]** While our mRNASeq data do not show an increase in mTOR levels, SYS derived fibroblasts have been reported to show an increase in mTOR expression and activity.**[63]**

**Figure 4.**
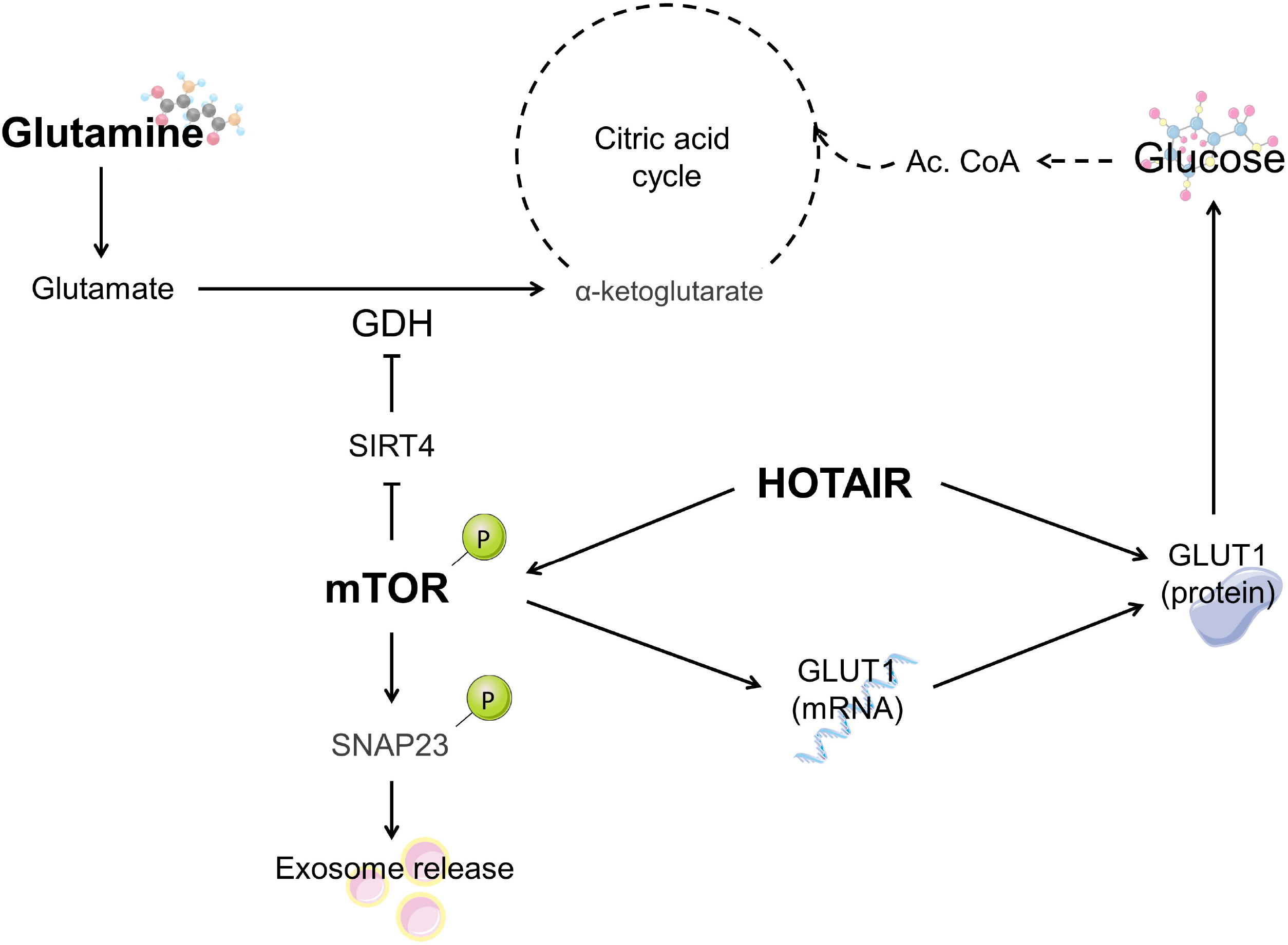
Schematization of the relationship between the different biomarkers identified in the SYS fibroblasts. The increased levels of HOTAIR could lead to the upregulation of mTOR previously seen in the literature as well as the deregulation of Glucose uptake through GLUT1.

Endocrine and metabolic alterations in PWS have been widely studied.**[64]** However, little is known regarding metabolic impairment in SYS. A study including five female and four male SYS patients showed that they may present some but not all the endocrine alterations also observed in PWS: Patients benefit from growth hormone therapy, show increased ghrelin levels, and present a high risk of developing diabetes mellitus.**[10]** A review on the literature on endocrine abnormalities in MAGEL2-related syndromes**[26]** showed that the most common hormonal alterations involve growth hormone (GH), thyroid stimulating hormone (TSH), adrenocorticotropic hormone (ACTH), antidiuretic hormone (ADH) and gonadotropins, probably caused by hypothalamic impairment.

The targeted metabolomic analysis of SYS and PWS derived fibroblasts was aimed at finding biomarkers that assess the lysosomal, peroxisomal, mitochondrial, and cytosolic metabolic pathways. The most relevant finding was a significant reduction in glutamine levels in SYS fibroblasts compared to the other two groups, whereas glutamate levels remained unchanged. Glutamine supplies nitrogen and carbon for biosynthetic reactions in rapidly proliferating cells,**[65]** but in many contexts its key role relies on providing glutamate (**Figure 4**), which has a wider range of metabolic functions than glutamine itself. An example of the relevant functions of glutamine and glutamate is the glutamate/GABA-glutamine cycle in neurons.**[66]** In fact, it has been stated that alterations of the GABAergic system may play an important role in aspects of the pathophysiology of PWS.**[67]** A hypothesis to explain the decrease in glutamine levels in SYS fibroblasts could be a dysregulation in the retrieval and recycling of a glutamine transporter, such as SLC1A5, whose retrieval and recycling is promoted by the retromer and whose degradation is enhanced upon retromer knockout.**[68]** Another explanation could be related to the hyperactivation of the mTOR pathway, as mTOR plays a role in the glutamine metabolism by increasing glutamate dehydrogenase activity through the inhibition of SIRT4 (**Figure 4**). **[69]** The hyperactivation of mTOR could drive a glutamate depletion which would be compensated by an increase in glutamine deamidation.

In conclusion, our results support the hypothesis that the SYS-specific phenotype might be explained by a gain-of-function effect of the truncated protein, rather than by a dose-reduction situation. This is supported by the subtle changes in gene expression patterns and metabolite levels observed in fibroblasts, which suggest novel effects for the truncated protein, which warrant further exploration. In addition, we provide a comprehensive phenotypical delineation of the syndrome and a standardised set of guidelines aimed at the improvement of clinical management of SYS patients. Finally, we propose to improve SYS diagnosis by testing the pathogenicity of doubtful missense mutations through the *in vitro* overexpression system.

## MATERIALS & METHODS

An extended version of the methodology is included as supplementary material. Briefly, we performed a systematic review of the literature indexed in PubMed from the date of the first clinical description of pathology associated with variants in *MAGEL2* until February 2022 using the terms: ‘*MAGEL2’*, ‘*SYS’*, ‘Schaaf-Yang syndrome’. All mutations have been referenced to the *MAGEL2* hg38 main transcript NM_019066.5. Fibroblasts were obtained from skin biopsies of seven SYS individuals, nine PWS patients and eleven controls (**Supplementary Table 5**). All cell cultures (fibroblasts, HEK293T, HeLa and SAOS-2) were cultured in standard cell culture conditions. The cDNA sequences of interest were cloned in the mammalian expression vector pcDNA™3.1^(+)^ (Invitrogen, ThermoFisher Scientific) including a HA tag and transfected into the different cell lines. Cells were treated with MG132, bafilomycin or cycloheximide. Whole RNA was extracted from fibroblasts and RNA-Seq was performed by LEXOGEN, Inc. and analysed with the ExpHunter Suite. **[70]** Expression levels of four selected genes were analysed by qPCR. Protein was extracted using RIPA buffer, resolved by SDS-PAGE and immunoblotted following standard biochemical techniques. For Immunocytochemistry, cells were fixed in 4% PFA, permeabilized and blocked. Coverslips were incubated with anti-HA primary antibody and DAPI. For ELISA quantification, the supernatant of the collected media (without FBS) was obtained and analysed by the Amyloid-beta (1-40) High Sensitive ELISA or the Amyloid-beta (1-42) High Sensitive ELISA (IBL International GmbH). For metabolomics, amino acids were analysed using ultra-high-performance liquid chromatography–tandem mass spectrometry (UPLC-MS/MS) and organic acids with gas chromatography-mass spectrometry (GC-MS). Statistical analysis was performed using R-Studio. Differences were considered significant if P < 0.05.

All skin donors or their legal representative gave their written informed consent. Their samples and data were obtained in accordance with the Helsinki Declaration of 1964, as revised in October 2013 (Fortaleza, Brazil). The study was approved by the Institutional Review Board (IRB00003099) of the Bioethical Commission of the University of Barcelona (October 5, 2020) and Hospital Sant Joan de Déu (PIC-111-19).

## Data Availability

Data are available upon reasonable request

## ACKNOWLEDGMENTS

We would like to thank patient’s associations AESYS and PWS-Catalunya and all the families who generously donated samples. We thank Mrs. Monica Cozar for her technical assistance.

## DATA AVAILABILITY

Data are available upon reasonable request.

## PATIENT CONSENT FOR PUBLICATION

All fibroblasts’ donors, or their parental/guardian, provided informed consent.

## ETHICS APPROVAL

Institutional ethics approval was obtained from the Ethics Committee of the Universitat de Barcelona, IRB00003099.

