## Supplemental material for "Advancing in Schaaf-Yang syndrome pathophysiology: from bedside to subcellular analyses of truncated MAGEL2"

**SUPPLEMENTARY FIGURES:**

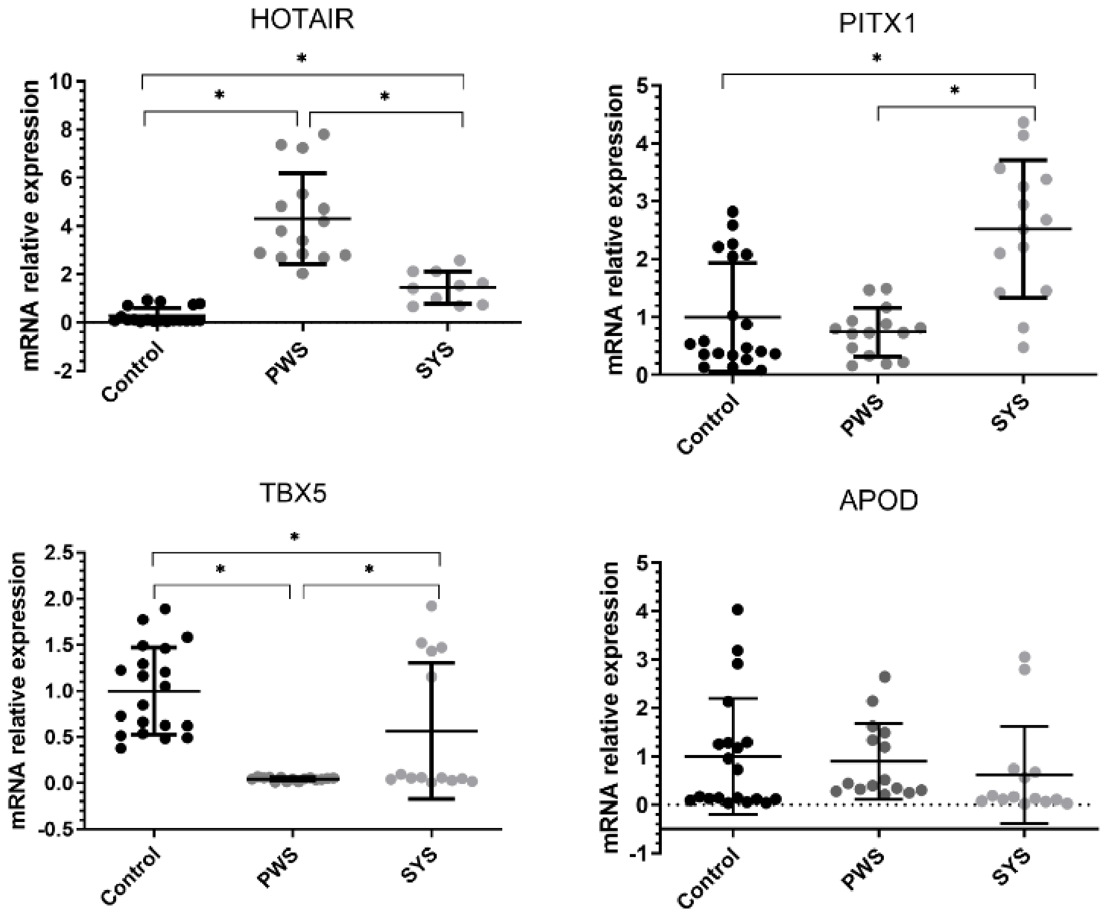


**Supplementary Figure 1:** qPCR analysis of *HOTAIR, PITX1, TBX5* and *APOD* in 5 SYS patients, 5 PWS patients and 7 control individuals. Values from 3 replicates have been normalised to the mean of the control group. Horizontal lines represent mean values and error bars represent the standard deviation. Statistical analyses were performed using One-Way ANOVA and Tukey's multiple comparisons test in GraphPad Prism.*$p< 0.001$.

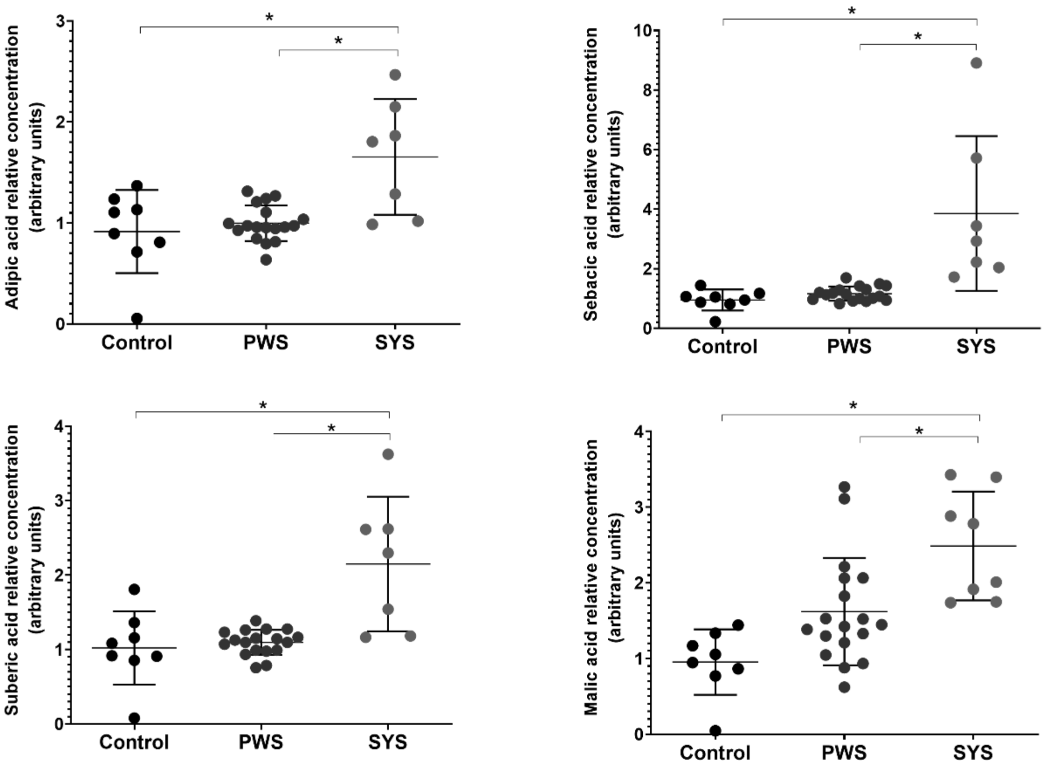


**Supplementary Figure 2:** Targeted metabolomics in fibroblasts. Adipic, Suberic, Sebacic and Malic acid are significantly increased in SYS derived fibroblasts compared to controls and PWS derived fibroblasts. (SYS: n=6; PWS: n=9; control: n=6). Values from 2 replicates have been normalised to the mean of the control group. Horizontal lines represent mean values and error bars represent the standard deviation. Statistical analyses were performed using One-Way ANOVA and Tukey's multiple comparisons test in GraphPad Prism.*p< 0.001.

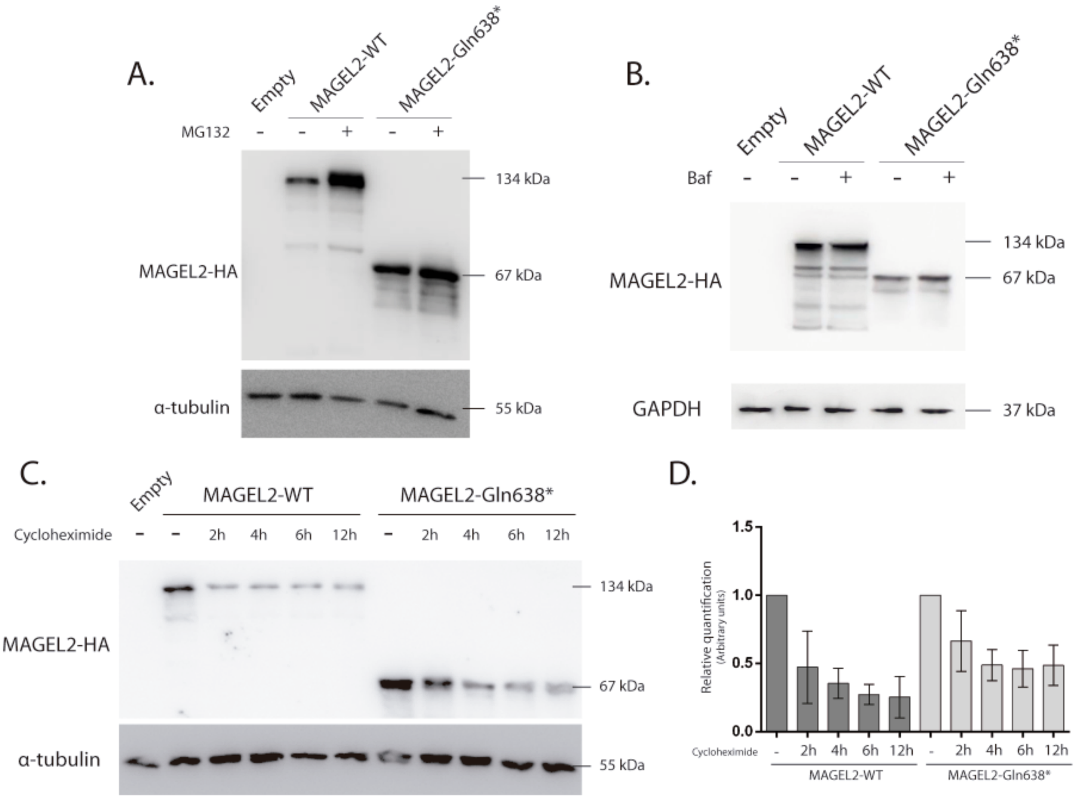


**Supplementary Figure 3: Degradation rate and stability of MAGEL2-WT and MAGEL2-Gln638* proteins.** A) Representative Western Blot of heterologously expressed MAGEL2 proteins with or without MG132 treatment (10µM for 16 hours) (n=3). B) Representative Western Blot of heterologously expressed MAGEL2 proteins with or without Baf treatment (1µM for 16 hours) (n=3). C) Representative Western Blot of heterologously expressed MAGEL2 proteins with or without Cx treatment (150µM) during 2, 4, 6 or 12 hours (n=3). D) Quantification of three independent experiments normalised to the expression levels of MAGEL2-WT or MAGEL2-Gln638* without Cx treatment.

### SUPPLEMENTARY MATERIAL AND METHODS:

**Literature review:** A Cochrane Library search and a PubMed database search was performed from the date of the first clinical description of pathology associated with variants in *MAGEL2* till May 31, 2022, using the following terms: '*MAGEL2'*, '*SYS'*, 'Schaaf-Yang syndrome', and different combinations of these terms. Concerning the data extraction, it included first author, publication year, molecular data (identified mutation and residue changes, *de novo* or inherited condition), pregnancy and perinatal information, multiorganic clinical data, complementary exams information, neuroimaging, and age and cause of death. Unfortunately, in some clinical series, much clinical information is not detailed.

**RNA-sequencing and data analysis:** Whole RNA was extracted from fibroblasts using the High Pure RNA Isolation Kit (Roche). RNA-Seq was performed by LEXOGEN, Inc. using the QuantSeq 3' messenger RNA (mRNA)-Seq FWD kit for library preparation. ExpHunter Suite **[1]** was used to analyse the expression data and perform functional enrichment analyses. The differential expressed genes (DEGs) are obtained using an adjusted p-value of 0.05, a minimum log fold change of 1 and Deseq2 and EdgeR packages give as DEG the analyzed gene. These DEGs are used as input for a functional enrichment analyses using REACTOME database and an adjusted p-value of 0.05.

**qPCR analysis of selected genes:** Whole RNA was extracted from fibroblasts using the High Pure RNA Isolation Kit (Roche). The extracted RNA was reverse transcripted using the High Capacity cDNA reverse transcription kit (Applied Biosystems) following the manufacturer's instructions. qPCR was performed using LightCycler® 480 SYBR Green I Master (04887352001, Roche). The human *GAPDH* gene served as a housekeeping gene and the efficiency of each reaction was calculated according to the standard curve. Melting-curves were conducted at the end of amplification to ensure data quality. Each sample was performed in triplicate. Primer sequences are available on demand.

**Cell culture:** Fibroblasts were obtained from skin biopsies of seven SYS patients, nine PWS patients and eleven controls (**Supplementary Table 6**). Corresponding informed consent and institutional ethics approval were obtained (Ethics Committee of the Universitat de Barcelona, IRB00003099). Fibroblasts were cultured in Dulbecco's Modified Eagle Medium (DMEM) (Sigma-Aldrich, Merck) supplemented with 10% Foetal Bovine Serum (FBS) (Gibco, LifeTechnologies), 1% Penicillin–Streptomycin (Gibco, LifeTechnologies) and 1% GlutaMAX (Gibco, LifeTechnologies). Conditioned medium for ELISA was only supplemented with 1% Penicillin–Streptomycin (Gibco, LifeTechnologies) and was collected after 72 h. HEK293T, HeLa and SAOS-2 cells were cultured in DMEM (Sigma-Aldrich, Merck) supplemented with 10% FBS (Gibco, LifeTechnologies) and 1% Penicillin–Streptomycin (Gibco, LifeTechnologies). The cDNA sequences of interest (ENST00000650528.1) were cloned in the mammalian expression vector pcDNA™3.1$^{(+)}$ (Invitrogen, ThermoFisher Scientific) including a hemagglutinin (HA) tag at the C-terminal end (courtesy of the CRG). The vectors were transfected into 60% confluent HEK293T, HeLa and SAOS-2 cells using Lipofectamine™ 3000 (Invitrogen, ThermoFisher Scientific) and Opti-MEM™ (Gibco, LifeTechnologies). Cells were treated with 10 µmol/L MG132 (Calbiochem) or 1 µmol/L bafilomycin (Sigma-

Aldrich, Merck) for 16 h or 150 µmol/L cycloheximide (Sigma-Aldrich, Merck) for 2, 4, 8 or 12 h prior to total protein extraction.

**Protein extraction and western blotting:** Total protein extraction was performed using RIPA buffer supplemented with protease inhibitors (04693159001, Roche, Merck) and N-ethylmaleimide (Sigma-Aldrich, Merck) and quantified using the Pierce™ BCA Protein Assay kit (ThermoFisher Scientific). Twenty µg of total protein per lane were run in 10% acrylamide/bis-acrylamide gels, transferred to a PVDF membrane (Millipore, Merck) and blocked with 5% skimmed milk in TBS-Tween 1X. Different antibodies were used: Anti-HA tag (ab18181, Abcam), anti-α-Tubulin (T5168, Sigma-Aldrich), anti-GAPDH (sc-47724, Santa Cruz Biotechnology) and anti-Mouse IgG (Fc specific)–Peroxidase (A0168, Sigma-Aldrich). Results were visualised using the LAS-4000 Luminescent Image Analyzer (Fujifilm) and the Luminata™ Forte Western HRP Substrate (WBLUF0100, Millipore). The detected bands were quantified using ImageJ.**[2]**

**Immunocytochemistry:** Cells were fixed in 4% Paraformaldehyde (PFA), permeabilized with 0,1 mol/L glycine and 0,1% Triton X-100 in PBS and blocked with 0,3 mol/L glycine, 0,05% Triton X-100 and 10% Normal Donkey Serum (#S30-100M, Merck Millipore) in PBS. Coverslips were incubated with anti-HA primary antibody (ab18181, Abcam), Donkey anti-Mouse Cy2 antibody (#715-225-150, Jackson Immunoresearch) and DAPI (#D1306, Invitrogen) and mounted with MOWIOL (#475904, Millipore). Images were acquired using a Zeiss confocal microscope LSM 880 and analysed with ImageJ. **[2]**

**ELISA analysis:** The supernatant of the conditioned medium was obtained after short centrifugation. Total protein concentration was quantified using the Pierce™ BCA Protein Assay kit (ThermoFisher Scientific) and $A\beta_{1\text{-}40}$ and $A\beta_{1\text{-}42}$ amyloid peptide using the Amyloid-beta (1-40) High Sensitive ELISA and Amyloid-beta (1-42) High Sensitive ELISA (IBL International GmbH). Statistical analyses were performed using One-Way ANOVA and Tukey's multiple comparisons test in GraphPad Prism.

**Metabolomics analyses:** Amino acid analyses were performed using a ultra-high-performance liquid chromatography–tandem mass spectrometry (UPLC-MS/MS) method, as previously reported.**[3]** Gas chromatography-mass spectrometry (GC-MS) analyses were performed to identify different organic acids, as described in **[4, 5]**. Statistical analyses were performed using One-Way ANOVA and Tukey's multiple comparisons test in RStudio (https://www.rstudio.com/).

**Ethical issues:** For the patients whose fibroblasts have been studied, parents gave their written informed consent. Their samples and data were obtained in accordance with the Helsinki Declaration of 1964, as revised in October 2013 (Fortaleza, Brazil). The study was approved by the Institutional Review Board (IRB00003099) of the Bioethical Commission of the University of Barcelona (October 5, 2020) and Hospital Sant Joan de Déu (PIC-111-19).
